# Integrating structured and unstructured EHR data to characterize social and behavioral factors associated with frequent emergency department use among patients with cardiovascular disease

**DOI:** 10.64898/2026.09.11.26362813

**Authors:** Tai Metzger, Shane Morrell, Sujoy Roy, Joshua Neumann, Lavi Singh, Payal Shah, Brian Felice, David Berger, Ramin Homayouni

## Abstract

**Objective:** The goal was to integrate structured and unstructured data from the electronic health record (EHR) to identify social and behavioral determinants of health (SBDH) factors and evaluate their association with high emergency department (ED) utilization among cardiovascular disease (CVD) patients.

**Methods:** A custom natural language processing (NLP) algorithm was developed to extract 17 SBDH domains from clinical notes, combined with ICD-10 Z codes and screening questionnaires from the EHR. The study included patients aged 18-64 with a history of CVD. Multivariable logistic regression was used to identify associations between SBDH and high ED utilization (≥ 5 visits) after adjusting for age, sex, race, insurance class, and chronic conditions.

**Results:** Use of only ICD-10 codes and screening questionnaires identified patients in only a subset of the 17 SBDH domains, whereas extraction from clinical notes identified patients across all domains and increased prevalence estimates for every domain. Among 4,844 patients with CVD, 526 (10.9%) were frequent ED users. After adjustment, patients with inadequate support system (OR= 2.8), opioid abuse (OR=2.47), inadequate insurance (OR=2.42), alcohol abuse (OR=2.28), medication affordability concerns (OR=2.28), unreliable transportation (OR=1.82), financial strain (OR=1.69), and depression (OR=1.42) were significantly more likely to have frequent ED utilization.

**Conclusion:** NLP can efficiently and more comprehensively identify socioeconomic risk factors from the EHR. Substance use, financial strain, inadequate social support, and other socioeconomic challenges may be linked to increased ED visits in patients with CVD.

## INTRODUCTION

### Background

Social and behavioral determinants of health (SBDH) play a role in a variety of patient health outcomes and healthcare utilization patterns.^1–9^ This is particularly relevant in patients with cardiovascular disease (CVD), a leading cause of death and emergency department (ED) visits in the United States.^1^ Electronic health records (EHR) contain valuable information regarding patient SBDH, but difficulties in extracting these data have limited their use in research.^10^ Recent advances in Natural Language Processing (NLP) approaches have enabled more comprehensive identification of SBDH evidence documented in clinical notes. For instance, an NLP algorithm was shown to identify seven social determinants of health domains from the EHR of Alzheimer’s patients.^11^ Large language models (LLMs) have also demonstrated the ability to identify SBDH evidence from clinical documentation, although application of LLMs to large amounts of clinical notes is prohibitively costly and time-consuming.^12^

### Importance

ED overutilization is a significant burden on the healthcare system, increasing costs and decreasing quality of care.^13,14^Previous studies have demonstrated a significant role of social factors in healthcare utilization. Inadequate health literacy, depression, impaired cognitive functioning, public insurance, lower education, lower household income, and more missed appointments were all found to be predictors of ED visits.^13^ Although some evidence suggests that frequent ED users have lower socioeconomic status (SES) and multiple mental and physical health conditions, more research is needed to better understand the factors that lead to chronic frequent ED use.^14^ A 2021 observational study found that lack of social support and broader biopsychosocial frailty were associated with increased ED utilization among adults over the age of 65.^15^ For children in fragile families, hospitalization history, utilization of the ED by their primary caregiver, and outpatient visits were associated with increased ED use.^16^ Violence, housing instability, employment or financial problems, social or family problems, lack of access to care or transportation, and nonspecific psychosocial needs were also positively associated with ED use.^17^ The ED is an especially challenging place to apply knowledge about SBDH to improve patient care because of the short amount of time ED physicians have with patients and the inability to focus on these factors during an ED visit.^18^

### Goals of This Investigation

While SBDH associated with high ED utilization have been extensively studied, these associations remain poorly characterized among patients with chronic cardiovascular disease (CVD). It is important to study this population because patients with different chronic conditions may have higher rates of ED utilization for distinct reasons. SBDH factors, including educational attainment, neighborhood/physical environment, economic stability, and race/ethnicity, have been linked to poorer cardiovascular disease outcomes, including stroke, myocardial infarction (MI), coronary heart disease, heart failure, and mortality.^19, 20^ Addressing SBDH factors in the ED may impact post-discharge cardiovascular events and provide guidance for health policies aimed at reducing ED overuse.^21^

## METHODS

### Study Design & Population

We performed a retrospective chart review of patients who utilized the Corewell Health William Beaumont University Hospital ED from September 1, 2022 to August 31, 2023. The study protocol was determined to be exempt from human subjects research requirements, with a waiver of informed consent, by the Corewell Health Institutional Review Board.

The study population included adult patients aged 18-64 years at the time of service and a history of one or more of the following cardiovascular diseases based on ICD-10 codes in the medical history or problem list: acute myocardial infarction (MI), atrial fibrillation (AF), non-ischemic heart disease, or ischemic heart disease. Patients 65 and older were excluded due to higher age-related chronic disease burden that may contribute to frequent ED utilization.

### SBDH algorithm development and evaluation

We developed an algorithm to extract SBDH from multiple sources within the EHR and generate patient-level reports summarizing the SBDH evidence for each patient (see pseudocode in Supplementary Materials). The sources of SBDH included ICD-10 z-codes in any prior billing encounter, medical history, or problem list. In addition, SBDH was extracted from responses to the standard screening forms in the EHR classified as medium or high risk.

Lastly, SBDH were extracted from all unstructured notes attributed to the patient within the study period, except for ‘patient instructions’ and ‘discharge instruction’ notes. The 17 SBDH domains included: food insecurity, housing insecurity, financial strain, unreliable transportation, alcohol abuse, opioid abuse, cocaine use, nicotine dependence, domestic violence/intimate partner violence, social isolation, inadequate support system, uninsured or underinsured status, medication affordability concerns, postpartum depression, depression, lack of physical activity, and stress (Supplementary Table S1).

We manually constructed Regular Expression (RegEx) patterns for each SBDH domain and used Linux grep function to search the clinical notes. For example, the RegEx pattern “living in a (?|camper|tent|car|shelter|van|truck)” was used to identify documentation suggestive of housing insecurity (Supplementary Table S1). The complete set of RegEx patterns is available upon request.

A Python-based workflow was developed to orchestrate the extraction, processing, and integration of SBDH data across structured and unstructured EHR sources. For each patient, the algorithm integrated evidence identified from ICD-10 codes, screening questionnaire results, and RegEx-based searches of clinical notes. A patient-level SBDH report was then generated, with each row representing an individual patient and columns representing evidence corresponding to each of the 17 SBDH domains. The SBDH evidence generated by the algorithm were subsequently transformed into binary variables and evaluated against a manually established SBDH gold standard (see below). Algorithm performance was evaluated using precision, recall, and F1 score. Precision represents the proportion of algorithm-identified positive cases that were true positives, whereas recall represents the proportion of all true positive cases that were correctly identified by the algorithm. The F1 score represents the harmonic mean of precision and recall, providing a combined measure of both metrics.

### Gold Standard Development

To generate a gold-standard dataset, we randomly selected 20 patients per each SBDH domain who were aged 18 years or older, had an ED encounter in 2023, and had at least one domain-relevant ICD-10 billing code associated with the encounter. All clinical notes linked to the qualifying ED encounters were extracted. Using the same age and encounter criteria, we also randomly sampled patients without an SBDH-related ICD-10 code and extracted all clinical notes associated with their qualifying encounters. This sampling strategy initially identified 310 patients with SBDH-related ICD-10 codes and 250 patients without SBDH-related ICD-10 codes. After excluding patients with no clinical notes, the final consolidated dataset included 548 patients and 49,285 clinical notes. For each patient, all notes were concatenated into a single document.

Each patient-level document was independently reviewed by four reviewers (TM, JN, LS, and RH) using the SBDH domain definitions provided in Supplementary Table S2. For each patient and SBDH domain, the gold-standard classification was determined by majority agreement. A domain was classified as present when at least three of the four reviewers agreed that the available clinical documentation supported its presence. The pairwise agreement between evaluators was determined using Cohen’s kappa (Supplementary Table S3).

## Statistical Analysis

The primary outcome was ≥ 5 ED visits during the study period, although there is no consensus definition of high ED utilization.^14^ For each individual in the study cohort, the following variables were extracted from the EHR: age, sex, race, 30 chronic conditions (Supplementary Material, Table S4),^22^ insurance plan type (Medicaid, Medicare, commercial, or other), primary ED billing diagnoses, frequency of ED encounters during the study period, and the 17 SBDH domains as described above.

For statistical analyses, categorical variables were compared using the chi-square test or Fisher exact test as appropriate, and continuous variables were compared using Kruskal-Wallis test. Two multivariable regression models were evaluated. Model 1 included demographic characteristics, insurance plan type, and SBDH domains, whereas Model 2 additionally included chronic conditions. Covariates were retained using backward stepwise selection with a significance threshold of P < 0.05. Statistical significance was defined as P < 0.05 for all analyses.

## RESULTS

### Construction and evaluation of the gold standard set

SBDH domain classifications of clinical notes from 548 randomly selected patients were determined by majority vote among four independent evaluators. The resulting domain counts and mean pairwise Cohen’s kappa values across evaluators were as follows (Supplementary Table S3): food insecurity, 21 (κ=0.815); housing insecurity, 77 (κ=0.895); financial strain, 99 (κ=0.911); unreliable transportation, 64 (κ=0.836); alcohol abuse, 114 (κ=0.832); opioid abuse, 64 (κ=0.862); cocaine use, 63 (κ=0.985); nicotine dependence, 232 (κ=0.978); domestic violence, 33 (κ=0.818); isolation, 52 (κ=0.579); inadequate social support, 80 (κ=0.682); uninsured or under-insured status, 66 (κ=0.911); medication affordability concerns, 49 (κ=0.893); postpartum depression, 21 (κ=1.000); depression, 242 (κ=0.930); lack of physical activity, 52 (κ=0.975); and stress, 168 (κ=0.813). Overall, inter-rater agreement was high across most SBDH domains, with the strongest agreement observed for postpartum depression, cocaine use, nicotine dependence, and lack of physical activity, while agreement was comparatively lower for isolation and inadequate social support.

### Characteristics of study subjects

A total of 4,844 patients aged 18-64 years met the inclusion criteria for the study (Figure 1). Among these, 526 patients (10.9%) were identified as frequent ED users (≥5 ED visits), while 4,318 patients (89.1%) were classified as non-high utilizers (<5 visits/year).

**Figure 1.**
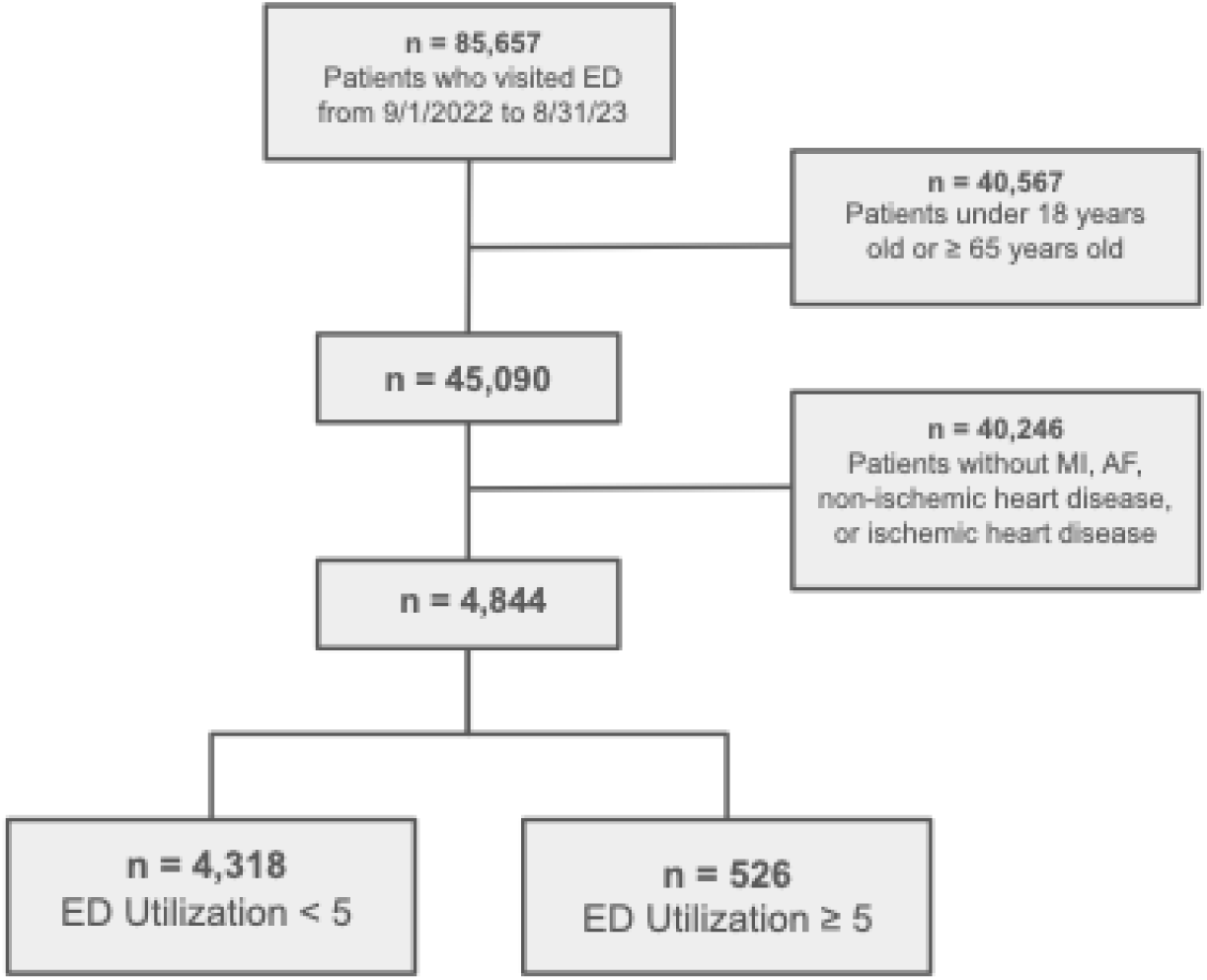
Study inclusion and exclusion workflow. A total of 85,657 patients visited the Corewell Health William Beaumont University Hospital from September 1, 2022 to August 31, 2023. Of these, 4,844 met inclusion criteria, with 4,318 in the non-high utilizers group and 526 in the frequent ED utilizers group.

### Extraction of SBDH from EHR

The algorithm was applied to the entire study cohort of ED patients with a history of CVD. Across the 17 SBDH domains, combining information from ICD codes, EHR screening forms, and clinical notes identified a higher prevalence than any individual data source alone (Table 1). Clinical notes captured the greatest proportion of cases for 12 domains, particularly alcohol abuse (10.96% vs 13.38% combined sources), opioid abuse (9.72% vs 12.63%), cocaine use (3.76% vs 3.96%), depression (26.05% vs 27.99%), and physical activity (7.97% vs 10.73%). In contrast, the EHR screening form was the predominant source for nicotine dependence, identifying 52.33% of patients compared with an overall prevalence of 53.74%, and also identified the highest proportion of patients with social isolation and stress. ICD codes contributed meaningfully to several substance-related domains, including alcohol abuse (8.79%), opioid abuse (5.35%), cocaine use (1.69%), and nicotine dependence (21.86%), but captured relatively few cases for several socioeconomic domains, such as food insecurity (0.02%), medication affordability concerns (0.08%), and uninsured/underinsured status (0.10%). The benefit of integrating sources was especially apparent for domains such as food insecurity (3.10% overall vs 0.02% ICD, 1.78% EHR screening form, and 1.38% notes), housing insecurity (4.07% vs 1.63%, 1.09%, and 1.90%, respectively), and financial strain (5.55% vs 2.00%, 2.19%, and 2.15%). Because patients could be identified by more than one source, the source-specific percentages are not additive; rather, the higher combined prevalence reflects the additional unique cases contributed by each source.

**Table 1.**
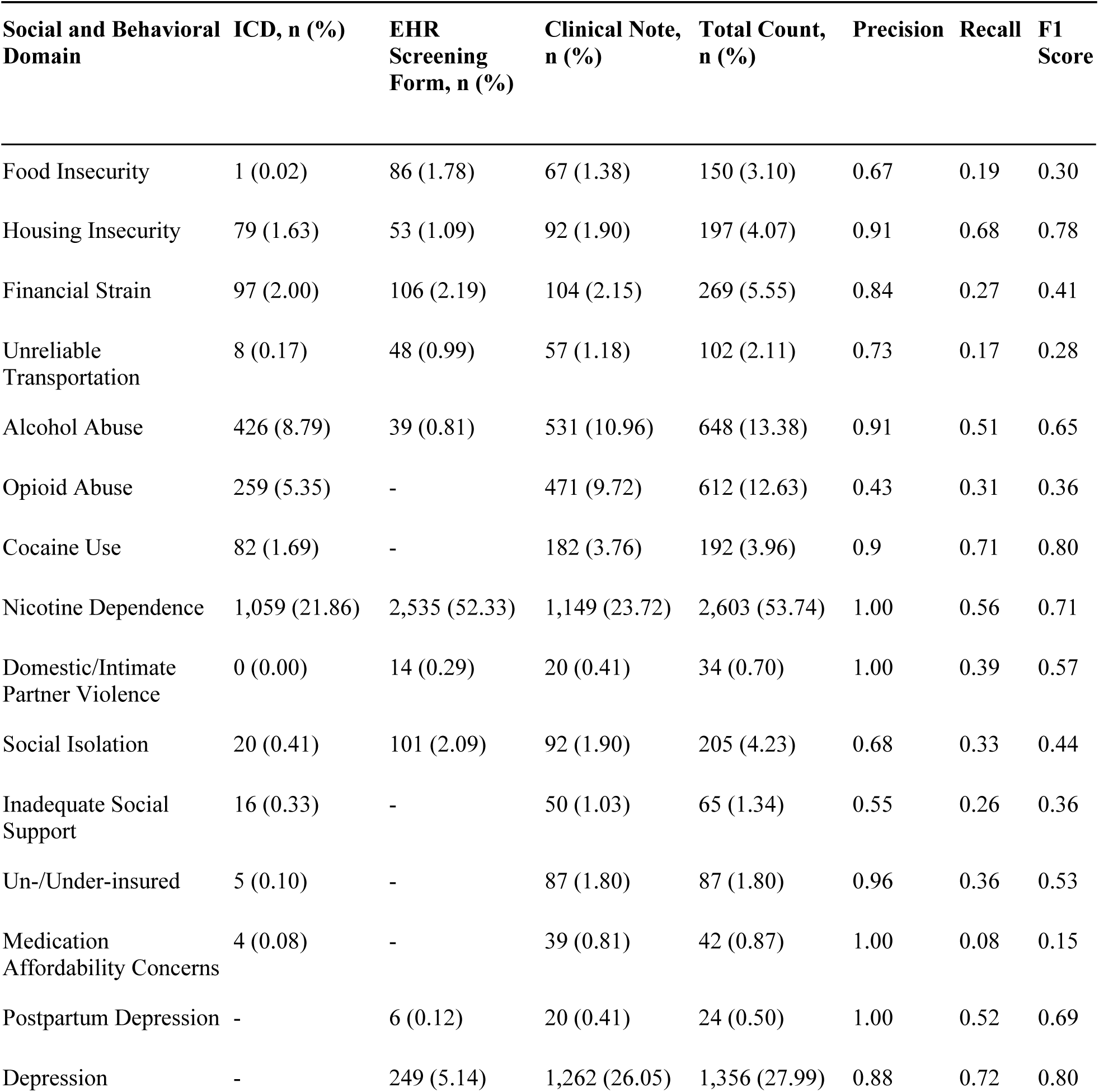
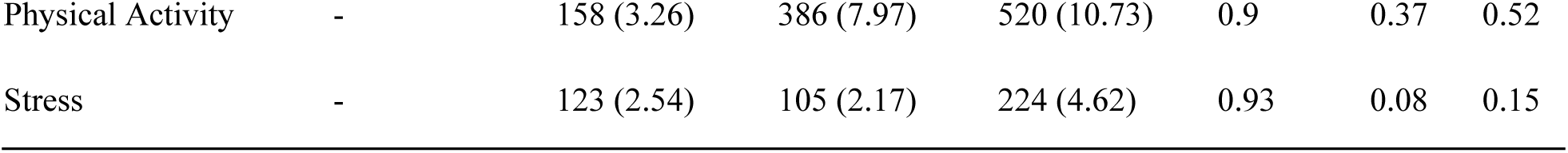
Prevalence and performance of SBDH domains detected from three different sources on the entire cohort (n=4,844). The precision, recall and F1 are derived from the total combined sources against a set of 548 curated gold-standard.

| Social and Behavioral Domain | ICD, n (%) | EHR Screening Form, n (%) | Clinical Note, n (%) | Total Count, n (%) | Precision | Recall | F1 Score |
| --- | --- | --- | --- | --- | --- | --- | --- |
| Food Insecurity | 1 (0.02) | 86 (1.78) | 67 (1.38) | 150 (3.10) | 0.67 | 0.19 | 0.30 |
| Housing Insecurity | 79 (1.63) | 53 (1.09) | 92 (1.90) | 197 (4.07) | 0.91 | 0.68 | 0.78 |
| Financial Strain | 97 (2.00) | 106 (2.19) | 104 (2.15) | 269 (5.55) | 0.84 | 0.27 | 0.41 |
| Unreliable Transportation | 8 (0.17) | 48 (0.99) | 57 (1.18) | 102 (2.11) | 0.73 | 0.17 | 0.28 |
| Alcohol Abuse | 426 (8.79) | 39 (0.81) | 531 (10.96) | 648 (13.38) | 0.91 | 0.51 | 0.65 |
| Opioid Abuse | 259 (5.35) | - | 471 (9.72) | 612 (12.63) | 0.43 | 0.31 | 0.36 |
| Cocaine Use | 82 (1.69) | - | 182 (3.76) | 192 (3.96) | 0.9 | 0.71 | 0.80 |
| Nicotine Dependence | 1,059 (21.86) | 2,535 (52.33) | 1,149 (23.72) | 2,603 (53.74) | 1.00 | 0.56 | 0.71 |
| Domestic/Intimate Partner Violence | 0 (0.00) | 14 (0.29) | 20 (0.41) | 34 (0.70) | 1.00 | 0.39 | 0.57 |
| Social Isolation | 20 (0.41) | 101 (2.09) | 92 (1.90) | 205 (4.23) | 0.68 | 0.33 | 0.44 |
| Inadequate Social Support | 16 (0.33) | - | 50 (1.03) | 65 (1.34) | 0.55 | 0.26 | 0.36 |
| Un-/Under-insured | 5 (0.10) | - | 87 (1.80) | 87 (1.80) | 0.96 | 0.36 | 0.53 |
| Medication Affordability Concerns | 4 (0.08) | - | 39 (0.81) | 42 (0.87) | 1.00 | 0.08 | 0.15 |
| Postpartum Depression | - | 6 (0.12) | 20 (0.41) | 24 (0.50) | 1.00 | 0.52 | 0.69 |
| Depression | - | 249 (5.14) | 1,262 (26.05) | 1,356 (27.99) | 0.88 | 0.72 | 0.80 |
| Physical Activity | - | 158 (3.26) | 386 (7.97) | 520 (10.73) | 0.9 | 0.37 | 0.52 |
| Stress | - | 123 (2.54) | 105 (2.17) | 224 (4.62) | 0.93 | 0.08 | 0.15 |

Based on the combined sources of SBDH, nicotine dependence was the most frequently identified domain (n = 2,603; 53.74%), followed by depression (n = 1,356; 27.99%), alcohol abuse (n = 648; 13.38%), opioid abuse (n = 612; 12.63%), and inadequate physical activity (n = 520; 10.73%). Other domains were less prevalent, including financial strain (5.55%), stress (4.62%), social isolation (4.23%), housing insecurity (4.07%), and cocaine use (3.96%),, with the remainder ranging from 0.5%-1.8%.

The NLP algorithm performance varied substantially across domains. Precision was generally high, reaching 1.00 for nicotine dependence, domestic/intimate partner violence, medication affordability concerns, and postpartum depression, and exceeding 0.90 for several additional domains. Recall was more variable and was generally lower than precision. The highest recall was observed for depression (0.72), cocaine use (0.71), and housing insecurity (0.68), whereas recall was particularly low for medication affordability concerns and stress (both 0.08), unreliable transportation (0.17), and food insecurity (0.19). The highest F1 scores were observed for cocaine use and depression (both 0.80), followed by housing insecurity (0.78), nicotine dependence (0.71), postpartum depression (0.69), and alcohol abuse (0.65). The lowest F1 scores were observed for medication affordability concerns and stress (both 0.15).

### Patient Characteristics and Univariate Analysis

Demographic and clinical characteristics differed significantly for high ED utilizers (Table 2). High utilizers had a lower median age of 54 years compared to non-high utilizers (median age 56, p<0.0001). A larger proportion of high utilizers were female (54.4%) compared to low utilizers (41.9%; p<0.0001). African American patients represented a higher percentage of high utilizers (46.0%) compared to low utilizers (34.0%, p<0.0001).

**Table 2.** Characteristics of CVD patients who frequently (≥5) visited the ED during a one-year period at Corewell Health Royal Oak Hospital. P-values were determined using Chi-Square or Fisher’s Exact test for categorical variables and Wilcoxon rank sum test for continuous variables.

| Variable | All | <5 ED visits | $\geq 5$ ED visits | P-Value |
| --- | --- | --- | --- | --- |
| **Sample, n (row %)** | 4844 | 4318 (89.1) | 526 (10.9) |  |
| **Age, median (IQR)** | 56 (11) | 56 (12) | 54 (12.5) | <.001 |
| **Female, n (column %)** | 2093 (43.2) | 1807 (41.9) | 286 (54.4) | <.001 |
| **Race, n (column %)** |  |  |  | <.001 |
| White/Caucasian | 2737 (56.6) | 2473 (57.4) | 264 (50.2) |  |
| Black/AA | 1706 (35.3) | 1464 (34.0) | 242 (46.01) |  |
| Asian | 107 (2.2) | 99 (2.3) | 8 (1.5) |  |
| Other | 288 (6.0) | 276 (6.4) | 12 (2.28) |  |
| **Insurance class, n (column %)** |  |  |  | <.001 |
| Commercial | 2297 (47.4) | 2180 (50.5) | 117 (22.2) |  |
| Medicaid | 1314 (27.1) | 1127 (26.1) | 187 (35.6) |  |
| Medicare | 1178 (24.3) | 957 (22.16) | 221 (42.0) |  |
| Other | 55 (1.1) | 54 | 1 (0.19) |  |
| **Number of SBDH, mean (std. dev)** | 1.51 (1.6) | 1.36 (1.4) | 2.75 (2.1) | <.001 |
| **Chronic Conditions, mean (std. dev)** | 5.8 (2.8) | 5.59 (2.7) | 7.61 (2.9) | <.001 |

Additionally, high utilizers were more likely to be covered by Medicaid (35.6%) or Medicare (42.0%), compared to low utilizers (26.1% and 22.16%, respectively; p<0.001). Also, high utilizers exhibited significantly greater burdens of chronic conditions (averaging 7.61 compared to 5.59 among low utilizers, p<0.0001). Similarly, high utilizers reported and average of 2.75 SBDH factors compared to 1.36 among non-high utilizers (p<0.0001).

### Multivariable Regression Analysis

Using multivariable logistic regression, we found several SBDH factors that were independently associated with frequent ED among patients with CVD. Model 1 adjusted for demographics and all 17 SBDH factors, while Model 2 additionally adjusted for 30 chronic conditions, with backward selection to keep only significantly associated variables (Table 3). Model fit improved after incorporating chronic conditions (R², 0.153 to 0.190). In Model 1, eight SBDH factors were found to be significantly associated with high ED utilization.

**Table 3.**
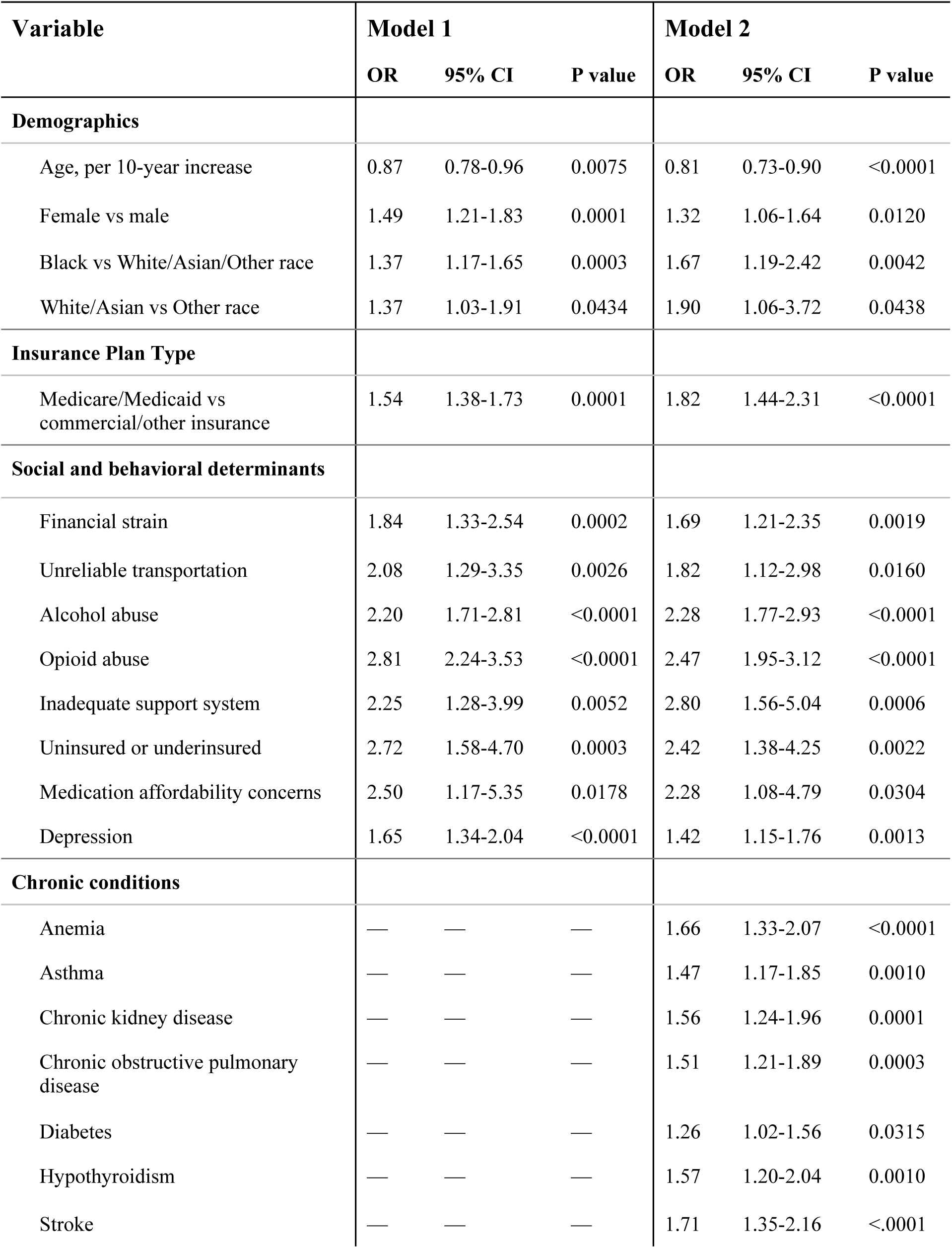
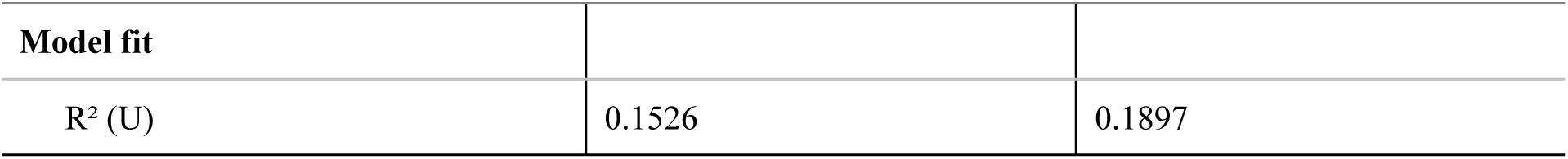
Multivariable logistic regression of factors associated with ≥ 5 ED visits.

| Variable | Model 1 |  |  | Model 2 |  |  |
| --- | --- | --- | --- | --- | --- | --- |
|  | OR | 95% CI | P value | OR | 95% CI | P value |
| **Demographics** |  |  |  |  |  |  |
| Age, per 10-year increase | 0.87 | 0.78-0.96 | 0.0075 | 0.81 | 0.73-0.90 | <0.0001 |
| Female vs male | 1.49 | 1.21-1.83 | 0.0001 | 1.32 | 1.06-1.64 | 0.0120 |
| Black vs White/Asian/Other race | 1.37 | 1.17-1.65 | 0.0003 | 1.67 | 1.19-2.42 | 0.0042 |
| White/Asian vs Other race | 1.37 | 1.03-1.91 | 0.0434 | 1.90 | 1.06-3.72 | 0.0438 |
| **Insurance Plan Type** |  |  |  |  |  |  |
| Medicare/Medicaid vs commercial/other insurance | 1.54 | 1.38-1.73 | 0.0001 | 1.82 | 1.44-2.31 | <0.0001 |
| **Social and behavioral determinants** |  |  |  |  |  |  |
| Financial strain | 1.84 | 1.33-2.54 | 0.0002 | 1.69 | 1.21-2.35 | 0.0019 |
| Unreliable transportation | 2.08 | 1.29-3.35 | 0.0026 | 1.82 | 1.12-2.98 | 0.0160 |
| Alcohol abuse | 2.20 | 1.71-2.81 | <0.0001 | 2.28 | 1.77-2.93 | <0.0001 |
| Opioid abuse | 2.81 | 2.24-3.53 | <0.0001 | 2.47 | 1.95-3.12 | <0.0001 |
| Inadequate support system | 2.25 | 1.28-3.99 | 0.0052 | 2.80 | 1.56-5.04 | 0.0006 |
| Uninsured or underinsured | 2.72 | 1.58-4.70 | 0.0003 | 2.42 | 1.38-4.25 | 0.0022 |
| Medication affordability concerns | 2.50 | 1.17-5.35 | 0.0178 | 2.28 | 1.08-4.79 | 0.0304 |
| Depression | 1.65 | 1.34-2.04 | <0.0001 | 1.42 | 1.15-1.76 | 0.0013 |
| **Chronic conditions** |  |  |  |  |  |  |
| Anemia | — | — | — | 1.66 | 1.33-2.07 | <0.0001 |
| Asthma | — | — | — | 1.47 | 1.17-1.85 | 0.0010 |
| Chronic kidney disease | — | — | — | 1.56 | 1.24-1.96 | 0.0001 |
| Chronic obstructive pulmonary disease | — | — | — | 1.51 | 1.21-1.89 | 0.0003 |
| Diabetes | — | — | — | 1.26 | 1.02-1.56 | 0.0315 |
| Hypothyroidism | — | — | — | 1.57 | 1.20-2.04 | 0.0010 |
| Stroke | — | — | — | 1.71 | 1.35-2.16 | <.0001 |

|  |  |  |
| --- | --- | --- |
| **Model fit** |  |  |
| R<sup>2</sup> (U) | 0.1526 | 0.1897 |

Importantly, even after adjustment for chronic conditions in Model 2, all eight SBDH factors remained significantly associated with frequent ED utilization as follows: inadequate support system (OR, 2.80; 95% CI, 1.56–5.04), followed by opioid abuse (OR, 2.47; 95% CI, 1.95– 3.12), uninsured or underinsured status (OR, 2.42; 95% CI, 1.38–4.25), alcohol abuse (OR, 2.28; 95% CI, 1.77–2.93), medication affordability concerns (OR, 2.28; 95% CI, 1.08–4.79), unreliable transportation (OR, 1.82; 95% CI, 1.12–2.98), financial strain (OR, 1.69; 95% CI, 1.21–2.35), and depression (OR, 1.42; 95% CI, 1.15–1.76).

## LIMITATIONS

This study has several limitations. First, generalizability may be limited because the data were derived from a single metropolitan hospital. Second, because ED visits at other hospitals were not available, ED visits are likely undercounted in our study. Third, given the retrospective observational design, causal relationships between the SBDH examined and increased ED utilization cannot be established. Fourth, there is a possibility of bias in estimating the SBDH prevalence of high-utilizers due to an increased likelihood of notations with more encounters in the health system. Fifth, although NLP enabled efficient extraction of SBDH information from clinical notes, some degree of algorithmic misclassification or incomplete capture of contextual language may have occurred. Sixth, ED utilization was assessed over a 1-year period, which may not fully reflect longer-term utilization patterns or temporal changes in SBDH. Longer follow-up may provide a more comprehensive assessment of utilization trajectories. Finally, potentially important factors, including neighborhood safety and health literacy, were not fully captured in the available EHR data.

## DISCUSSION

This study identified several key health-related social and behavioral needs that are strongly associated with frequent ED utilization among CVD patients. Notably, documented history of opioid and alcohol abuse, inadequate social support, being underinsured, and being unable to afford medications more than doubled the odds of high utilization after adjusting for demographic factors and chronic conditions. Importantly, SBDH factors had a higher effect (odds ratio) on high ED utilization than any of the significantly associated chronic conditions and demographic factors. These findings reinforce previous literature regarding the important role of SBDH in healthcare over-utilization beyond traditional clinical and demographic factors.^2–4, 14^

By integrating NLP to analyze clinical notes to detect SBDH needs, this study demonstrates an innovative approach to leveraging clinical notes for population health studies. Although some SBDH categories demonstrated relatively low F1 scores, the NLP algorithm provided value by identifying a number of SBDH factors that were not captured through the structured data sources. For example, substance use disorders are often not assessed in standard screening forms, whereas factors such as lack of physical activity or stress are not typically coded by diagnosis codes. These issues are exacerbated by the general underutilization of screening tools across large health systems.^23, 24^ Importantly, we demonstrated that each SBDH domain identified by the algorithm contributed additional cases to the overall prevalence, thus providing an avenue to supplement low screening rates across the health system. It is important to note that the lower F1 scores driven by missed cases would be expected to bias prevalence estimates toward underdetection rather than artificially inflate observed associations. Thus, the significant associations identified despite imperfect sensitivity of the algorithm may represent conservative estimates of the true relationships between SBDH and frequent ED utilization.

These findings support the utility of NLP-based approaches for supplementing structured EHR data and improving identification of socially relevant factors that may otherwise remain undocumented or inaccessible for large-scale research.

Adjustment for chronic disease burden further demonstrated the strength of the observed associations between SBDH and frequent ED utilization. In the initial regression model, before accounting for chronic conditions, inclusion of SBDH factors explained approximately 13% of the variation in the outcome (R² = 0.13). After chronic conditions were incorporated into the model, the proportion of explained variance increased to approximately 19% (R² = 0.19), indicating improved overall model performance while allowing the independent contribution of SBDH factors to be assessed beyond underlying disease burden. An explained variance of 19% is meaningful given the multifactorial nature of utilization.

Importantly, the persistence of SBDH associations after adjustment for chronic disease suggests that their relationship with ED utilization is independent of chronic disease burden and reinforces the importance of incorporating SBDH factors alongside traditional clinical variables in frequent ED utilization risk assessment.

Other studies have similarly reported associations between financial problems, depression, and inadequate social support with increased healthcare utilization.^17, 25^ In addition, substance use disorders, including opioid and alcohol abuse, were most strongly associated with frequent ED visits.^26^ Our study shows that these factors may also contribute to frequent ED utilization among CVD patients. Substance use may be a particularly important factor for over-utilization among CVD patients due to sympathetic activation, hypertension, arrhythmias, myocardial ischemia, and medication interactions. Specifically, opioids are known to negatively impact cardiovascular health through increased arterial stiffness, decreased heart rate variability, worsening coronary artery disease, and increased risk of atrial fibrillation.^27^

Cocaine causes increased cardiovascular disease through induction of hypertension, arrhythmias, and coronary vasospasm.^28^ These findings underscore the need for integrated care models that address both medical and social needs to reduce the burden of ED visits and improve overall health outcomes among CVD patients.

Future efforts should focus on implementing targeted interventions to address the identified SBDH factors among high-risk populations within the ED setting. For instance, integrating case management, social work, and community health worker support into CVD care models could help address financial strain, transportation barriers, and inadequate support systems. Substance use interventions, including medication-assisted treatment and counseling services, should also be prioritized to reduce the burden of opioid and alcohol abuse among CVD patients.

In summary, this study demonstrates that applying NLP for detection of SBDH from EHR offers a scalable, efficient strategy to uncover these frequently underreported risk factors. In addition, the study identified specific SBDH factors such as substance use, financial strain, inadequate social support, and transportation barriers that are strongly associated with frequent ED utilization among CVD patients. These findings highlight the need for targeted, multidisciplinary interventions that address both medical and social needs to reduce avoidable ED utilization.

## Data Availability

The data that support the findings of this study are available from the
corresponding author upon reasonable request.

