## Supplementary Material for "Integrating structured and unstructured EHR data to characterize social and behavioral factors associated with frequent emergency department use among patients with cardiovascular disease"

### Pseudocode for the SBDH Algorithm

---

```
Extract data from Epic Clarity:
    Export patient-level ICD diagnosis codes
    Export SBDH screening results and interpretations
    Export clinical note metadata and text for all note types

Aggregate clinical notes by patient:
    For each clinical note:
        If type not "Patient Instructions" or "Discharge Instructions":
            Add note text to the corresponding patient's document

Identify SBDH evidence for each predefined domain:
    For each SBDH domain:
        Identify ICD-based evidence:
            Load the predefined ICD codes associated with the domain
            For each ICD record:
                If the patient's ICD code matches a domain-specific code:
                    Mark the patient as having ICD-based evidence

        Identify screening-based evidence:
            For each SBDH screening record:
                If the domain-specific screening interpretation is
                "medium" or "high":
                    Mark the patient as having screening-based evidence

        Identify clinical note evidence:
            Load predefined regular-expression patterns for the domain
            Combine and compile the domain-specific patterns

            For each patient's aggregated clinical notes:
                Search the text using the domain-specific patterns
                If a match is identified:
                    Mark the patient as having NLP-based evidence
```

Record the matched text

Record ICD, screening, and NLP-based evidence in scorecard domain column

Export the final patient-level scorecard to a file

---

**Table S1.** ICD-10 Codes corresponding to SBDH domains along with example Regex patterns and corresponding clinical note matches.

<table>
<tr><th>Domain</th><th>ICD-10 Codes</th><th>Sample Regex Pattern</th><th>Example Match</th></tr>
<tr><td>Food Insecurity</td><td>Z59.4, Z59.41, Z59.48</td><td>((?:trouble with|has|significant|has some) food insecurity)</td><td>Trouble with food insecurity</td></tr>
<tr><td>Housing Insecurity</td><td>Z59.0, Z59.1, Z59.3, Z59.8, Z59.9, Z59.00, Z59.01, Z59.02, Z59.10, Z59.11, Z59.12, Z59.811, Z59.812, Z59.819</td><td>(type of home:\s{0,3}(?:[hm]otel|camper|tent|car|shelter))</td><td>Type of Home: shelter</td></tr>
<tr><td>Financial Strain</td><td>Z56.0, Z59.5, Z59.6, Z59.86, Z91.120</td><td>((?:occupation|employment):\s{0,3}unemployed)</td><td>Occupation: unemployed</td></tr>
<tr><td>Unreliable Transportation</td><td>Z75.3, Z59.82</td><td>((?:does not|doesn'?t|don'?t) have (?:a )?(?:vehicle|car\b|transportation))</td><td>Doesn’t have a vehicle</td></tr>
<tr><td>Alcohol Abuse</td><td>F10*</td><td>((?:alcohol|etoh) abuse:\s{0,3}yes)</td><td>Etoh abuse: yes</td></tr>
<tr><td>Opioid Abuse</td><td>F11*</td><td>((?:personal history of |history of |h/o |hx )?(?:opioid|heroin) {1,3}(?:abuse|dependence |addiction|overdose))</td><td>h/o heroin overdose</td></tr>
<tr><td>Cocaine Use</td><td>F14*</td><td>(cocaine) {0,3}\d{1,2}/\d{1,2}/\d{2,4} {0,3}pos</td><td>Cocaine 2/12/2023 pos</td></tr>
<tr><td>Nicotine Dependence</td><td>F17.210</td><td>((?:states|reports) daily tobacco)</td><td>Reports daily tobacco</td></tr>
<tr><td>Domestic Violence/Intimate Partner Violence</td><td>Z63.0, Z69.1, Z69.11</td><td>(Domestic (?:violence|abuse)(?: concerns)?[:\?]\s{0,3}yes )</td><td>Domestic abuse concerns? yes</td></tr>
<tr><td>Isolation</td><td>Z60.2, Z60.4, Z63.32, Z74.1, Z74.2</td><td>(Friends:\s{0,3}(?:no|Doesn't have friends|describes not having any friends))</td><td>Friends: no</td></tr>
<tr><td>Inadequate Support System</td><td>Z63.8, Z63.9</td><td>((?:does not|doesn'?t) have (?:a )?(?:good )?support system)</td><td>Doesn’t have a support system</td></tr>
<tr><td>Uninsured or Under-insured</td><td>Z59.7</td><td>((?:Didn't|Doesn't|does not|did not) have</td><td>Does not have insurance</td></tr>
</table>

<table>
<tr><td></td><td></td><td>insurance)</td><td></td></tr>
<tr><td>Medication Affordability Concerns</td><td>Z91.120</td><td>((?:cannot|can'?t|can not|could not|couldn'?t|not able to|won'?t be able to|unable to|inability to|lack of being able to)\b (?:pay for|afford) (?:the |his |her |their )?(?:medication|medicine|meds|prescription|rx|insulin))</td><td>Unable to pay for insulin</td></tr>
<tr><td>Postpartum Depression</td><td>F53*, O90.6, Z13.32</td><td>(?:she|patient) ((?:had|has|experienced) postpartum depression)</td><td>she has postpartum depression</td></tr>
<tr><td>Depression</td><td>F31.30, F31.31, F31.32, F31.4, F31.5, F31.60, F31.61, F31.62, F31.63, F31.64, F31.75, F31.76, F31.77, F31.78, F31.81, F32.0, F32.1, F32.2, F32.3, F32.4, F32.5, F32.9, F33.0, F33.1, F33.2, F33.3, F33.40, F33.41, F33.42, F33.8, F33.9, F34.1, F43.21, F43.23</td><td>(mood(?: and affect)?:\s{0,3}(?:depressed|mood is depressed))</td><td>Mood: depressed</td></tr>
<tr><td>Lack of Physical Activity</td><td>Z72.3</td><td>(physical activity:\s{0,3}(?:limited|none|nothing outside of work|inconsistent|not consistently|no consistent activity|minimal|restricted|wheelchair bound))</td><td>Physical activity: minimal</td></tr>
<tr><td>Stress</td><td>Z63.79, F43*</td><td>((?:has been|is) under a lot of stress)</td><td>Is under a lot of stress</td></tr>
</table>

**Table S2**. Definitions of SBDH domains used for manual evaluation.

| | |
|---|---|
| **Housing Insecurity** | Housing insecurity is present when the patient explicitly lacks stable, safe, adequate, or sustainable housing, including homelessness, temporary housing caused by lack of alternatives, risk of losing housing, frequent involuntary moves, or materially inadequate housing conditions. |
| **Food Insecurity** | Food insecurity is present when the record explicitly documents limited, uncertain, or inadequate access to sufficient food because of insufficient money or other household resources. |
| **Financial Insecurity** | Financial strain is present when the patient explicitly reports difficulty paying for basic necessities because available financial resources are insufficient. |
| **Unreliable Transportation** | Unreliable transportation is present when an explicit lack of dependable transportation prevents, delays, or threatens the patient's ability to obtain medical care, attend work or required meetings, or obtain necessities for daily living. |
| **Alcohol Misuse** | Alcohol misuse is present when the record explicitly documents problematic, hazardous, or harmful alcohol consumption, impaired control over drinking, alcohol-related impairment, or an active alcohol use disorder. |
| **Opioid Misuse** | Opioid misuse is present when the record explicitly documents nonmedical use of a prescription opioid, use in a manner other than prescribed, illicit opioid use, or an active opioid use disorder. |
| **Cocaine Use** | Cocaine use is present when the record explicitly documents current or recent use of cocaine or crack cocaine during the observation window. |
| **Nicotine Dependence** | Nicotine dependence is present when the record explicitly documents active nicotine dependence, tobacco use disorder, nicotine addiction, or another current dependence diagnosis involving a nicotine-containing product. |
| **Domestic Violence** | Domestic or intimate partner violence is present when the patient explicitly experiences physical violence, sexual violence, threats, stalking, psychological aggression, coercive control, or other abuse by a current or former intimate partner or, under a broader domestic-violence definition, a household or family member. |
| **Social Isolation** | Social isolation is present when the record explicitly documents an objective absence or marked scarcity of social relationships, social contact, or participation with other people. |
| **Inadequate Support** | Inadequate social support is present when the patient explicitly lacks sufficient emotional, informational, practical, caregiving, or day-to-day assistance to meet identified needs. |
| **Uninsured or Underinsured** | Uninsured means the patient has no active health care coverage. Underinsured means the patient has coverage, but the coverage is explicitly insufficient to obtain or afford needed health services. |
| **Medication Affordability** | Medication affordability concern is present when the record explicitly states that medication cost, copayment, deductible, or lack of prescription coverage makes it difficult or potentially impossible for the patient to |

| | |
|---|---|
| | obtain or take a prescribed medication. |
| **Postpartum Depression** | Postpartum depression is present when an active depressive disorder or positive postpartum depression screen is documented during the first 12 months after delivery, and the depression is explicitly identified as postpartum or perinatal, or is detected through a screening process specifically conducted for postpartum depression. |
| **Depression** | Depression is present when the record explicitly documents an active depressive disorder or a clinician affirmatively assesses the patient as currently experiencing depression or a depressive episode. |
| **Inadequate Physical Activity** | Lack of physical activity is present when the record explicitly documents physical inactivity or when quantified activity falls below the prespecified age-appropriate threshold. |
| **Stress** | The documentation explicitly states that the patient is experiencing psychological or emotional stress during the current encounter or predefined observation period. Qualifying evidence includes a patient statement of feeling stressed or under stress, a clinician assessment of current psychosocial or situational stress, or a positive response on a structured stress assessment. The presence of adverse social circumstances, anxiety, depression, insomnia, or other potentially related conditions does not qualify unless stress itself is explicitly documented. Mentions that are negated, historical and resolved, hypothetical, related to another person, or refer to physiological or technical uses of the word “stress” are excluded. |

**Table S3.** Pairwise and average Cohen's kappa values for each of the 17 SBDH domains.

| SBDH Domain | Count | Mean kappa (across pairs) |
|---|---|---|
| Food insecurity | 21 | 0.815 |
| Housing Insecurity | 77 | 0.895 |
| Financial Strain | 99 | 0.911 |
| Unreliable Transportation | 64 | 0.836 |
| Alcohol Abuse | 114 | 0.832 |
| Opioid Abuse | 64 | 0.862 |
| Cocaine Use | 63 | 0.985 |
| Nicotine Dependence | 232 | 0.978 |
| Domestic Violence | 33 | 0.818 |
| Isolation | 52 | 0.579 |
| Inadequate Social Support | 80 | 0.682 |
| Uninsured or Under-insured | 66 | 0.911 |
| Medication Affordability Concerns | 49 | 0.893 |
| Postpartum Depression | 21 | 1.000 |
| Depression | 242 | 0.930 |
| Lack of Physical Activity | 52 | 0.975 |
| Stress | 168 | 0.813 |

**Table S4.** Chronic Conditions Based on Centers for Medicare & Medicaid Services Database (version 2021).[22]

| Chronic disease condition | n | % of total population |
|---|---|---|
| Acute myocardial infarction | 1,313 | 27.1% |
| Alzheimer's disease | 7 | 0.1% |
| Anemia | 1,972 | 40.7% |
| Asthma | 975 | 20.1% |
| Atrial fibrillation | 1,333 | 27.5% |
| Hyperplasia | 284 | 5.9% |
| Breast cancer | 109 | 2.3% |
| Colorectal cancer | 63 | 1.3% |
| Endometrial cancer | 32 | 0.7% |
| Lung cancer | 56 | 1.2% |
| Prostate cancer | 65 | 1.3% |
| Urologic cancer | 42 | 0.9% |
| Cataract | 178 | 3.7% |
| Chronic kidney disease | 1,217 | 25.1% |
| COPD | 991 | 20.5% |
| Depressive disorder | 1,567 | 32.3% |
| Diabetes | 1,922 | 39.7% |
| Glaucoma | 129 | 2.7% |
| Non-ischemic heart disease | 1,916 | 39.6% |
| Hip/pelvic fracture | 57 | 1.2% |
| Hyperlipidemia | 3,115 | 64.3% |
| Hypertension | 3,885 | 80.2% |
| Hypothyroidism | 630 | 13.0% |

| | | |
|---|---|---|
| Ischemic heart disease | 3,048 | 62.9% |
| Non-Alzheimer dementia | 79 | 1.6% |
| Osteoporosis | 128 | 2.6% |
| Parkinson’s disease | 17 | 0.4% |
| Pneumonia | 939 | 19.4% |
| Rheumatoid arthritis | 1,308 | 27.0% |
| Stroke | 756 | 15.6% |
